# Learning Outcomes that Maximally Differentiate Psychiatric Treatments

**DOI:** 10.1101/2024.12.03.24318424

**Authors:** Eric V. Strobl, Semmie Kim

**Affiliations:** University of Pittsburgh

## Abstract

**Objective:** Matching each patient to the most effective treatment option(s) remains a challenging problem in psychiatry. Clinical rating scales often fail to differentiate between treatments because most treatments improve the scores of all individual items at only slightly varying degrees. As a result, nearly all clinical trials in psychiatry fail to differentiate between active treatments.

**Methods:** We introduce a new statistical technique called Supervised Varimax (SV) that corrects this problem by accurately detecting large treatment differences directly from original clinical trial data. The algorithm combines the individual items of a clinical rating scale that only *slightly* differ between treatments into a few scores that *greatly* differ between treatments. We applied SV to multi-center, double-blind and randomized clinical trials called CATIE and STAR*D which were long thought to identify few to no differential treatment effects.

**Results:** SV identified optimal outcomes harboring large differential treatment effects in Phase I of CATIE (absolute sum = 1.279, *p* = 0.002). Post-hoc analyses revealed that olanzapine is more effective than quetiapine and ziprasidone for hostility in chronic schizophrenia (difference = −0.284, *p*_*FWER*_ = 0.047; difference = −0.283, *p*_*FWER*_ = 0.048), and perphenazine is more effective than ziprasidone for emotional dysregulation (difference = −0.313, *p*_*FWER*_ = 0.020). SV also discovered that bupropion augmentation is more effective than buspirone augmentation for treatment-resistant depression with increased appetite from Level 2 of STAR*D (difference = −0.280, *p*_*FWER*_ = 0.003).

**Conclusions:** SV represents a powerful methodology that enables precision psychiatry from clinical trials by optimizing the outcome measures to differentiate between treatments.

## Introduction

A major goal of precision psychiatry is to match each patient to the most therapeutic treatment options(s) more precisely than the current standard of care [1]. However, nearly all randomized clinical trials (RCTs) in psychiatry fail to differentiate between the effects of active treatments on patient symptoms. For example, the CATIE trial compared the effects of multiple antipsychotics in chronic schizophrenia but could not pinpoint any pairwise differences between the effects of the antipsychotics on symptoms of schizophrenia [2]. Similarly, the STAR*D trial compared the effects of antidepressants and cognitive therapy in treatment-resistant depression but found little to no differential treatment effects [3, 4]. The original analyses of most RCTs unfortunately did not reveal differences between active treatments that would have otherwise enabled precision psychiatry.

Investigators have since re-analyzed datasets from the aforementioned and similar RCTs using progressively more complicated machine learning algorithms in order to reveal patient-specific differences in treatment effects. Many existing methods now utilize a multitude of baseline clinical and/or biological variables to predict treatment response (e.g., [5, 6, 7, 8, 9, 10, 11]). However, the ever-increasing number of variables and complexity of machine learning makes it more difficult to deploy, generalize, interpret, and maintain these algorithms in everyday clinical practice [12, 13]. Increasing data inputs and complexity can thus also diminish real-world usability.

Psychiatrists usually differentiate treatments by memorizing the unique effects of treatments on individual symptoms, or the individual items of a clinical rating scale in the context of research. For example, a psychiatrist may prescribe bupropion more often to patients with major depression, hypersomnia and weight gain because bupropion is activating and decreases appetite. We thus believe that many of the original RCTs did not identify differential treatment effects simply because the dependent variables, such as total severity scores, remission status and even validated sub-scales, amalgamate multiple symptoms into coarse outcome measures rather than differentiate treatments based on fine-grained analysis of all individual symptoms.

Differentiating treatments based on all individual symptoms is unfortunately not straightforward because treatments tend to diffusely affect nearly all items in a rating scale with weak signal and a large amount of noise. Searching for differential treatment effects among all individual items also yields equivocal results due to the burden of multiple hypothesis testing. We therefore design a new factor analysis technique called *Supervised Varimax* (SV) that takes the individual items from clinical rating scales and, unlike traditional approaches, identifies a few *optimal outcomes* that maximally differentiate between treatments. We can interpret the optimal outcomes as sub-scales or as representations of biopsychosocial processes that can best distinguish the effects of different treatments. We then estimate the effects of treatment on these optimal outcomes, rather than on the original dependent variables. We apply SV to CATIE and STAR*D in order to identify differential treatment effects on the optimal outcomes in chronic schizophrenia and treatment-resistant depression. We find many differential treatment effects that were undetectable in the original analyses. We finally glean simple, clinically-usable rules from the results that maximize treatment response using baseline data alone.

## Methods

### Clinical Trials

We analyzed a large-scale randomized clinical trial of schizophrenia called Clinical Antipsychotic Trials of Intervention Effectiveness (CATIE) [2], and another large-scale clinical trial of treatment-resistant depression called Sequenced Treatment Alternatives to Relieve Depression (STAR*D) [3, 4]. Both of these trials have been described in detail elsewhere. We included all patients with complete baseline data relevant to our analyses and always performed an intention-to-treat (last observation carried forward) analysis. We provide a brief summary of the components of the trials relevant to this paper below:

1. CATIE (ClinicalTrials.gov, NCT00014001, [2]) was a multi-center, double-blind, randomized clinical trial that compared atypical and typical antipsychotics in adult patients with chronic schizophrenia. We focus on Phase I of CATIE, where patients randomly received one of five treatment options: quetiapine, perphenazine, olanzapine, risperidone and ziprasidone.
2. STAR*D (ClinicalTrials.gov, NCT00021528, [3, 4]) was a multi-center, double-blind, randomized clinical trial that aimed to identify the most effective treatments for adult patients with depression whose symptoms did not remit after an initial prescription of citalopram. We focus on Level 2 of the STAR*D dataset, where participants received treatment only if they agreed to at least one of the following four options: medication switch, medication augmentation, cognitive therapy switch, and cognitive therapy augmentation. Patients then underwent randomization among the treatment options that they accepted. As a result, patients were strictly randomized only among (a) the medication switch options including bupropion, sertraline and venlafaxine, as well as (b) the medication augmentation options including buspirone augmentation and bupropion augmentation.

We downloaded the data of both studies from the National Institute of Mental Health (NIMH) Data Archive (https://nda.nih.gov/) with a limited access data use certificate awarded to author Eric V. Strobl.

### Original Outcome Measures

The CATIE study tracked antipsychotic response using the total score of the Positive and Negative Syndrome Scale (PANSS) [14]. The new algorithm described below takes individual items of a clinical rating scale as input. We thus input the values of all 30 individual items of the PANSS into the algorithm. On the other hand, the STAR*D trial tracked antidepressant response using the total score of the 16-item Quick Inventory of Depressive Symptomatology Self Report (QIDS-SR) score [15]. We thus input all of the individual 16 items of the QIDS-SR into our algorithm.

### Supervised Varimax

We provide a broad overview of the SV algorithm here and include a detailed technical description in Supplementary Materials 1.1-1.5. Briefly, the *original outcome* of most clinical trials corresponds to the remission status or the total severity score according to a clinical rating scale. SV instead considers the model in Figure 1 (a), where treatments ***T*** affect items on a rating scale ***Y*** by intermediately affecting a set of latent factors ***F*** representing unknown biopsychosocial processes. In general, treatments affect the factors ***F*** in complicated ways, and the factors ***F*** affect the items ***Y*** in complicated ways. This complexity diffusely distributes the treatment effects across many factors and items which makes it difficult to differentiate between treatments. SV organizes the complexity by learning an optimal set of factors ***F***^∗^ from the data such that the effects from treatments to ***F***^∗^ are maximally different and simple (Figure 1 (b)). For example, treatment *T*_1_ affects all factors ***F*** in Figure 1 (b) but only affects one optimal factor 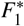 in Figure 1 (b). The optimized factors ***F***^∗^, which we also call *optimal outcomes*, thus now differentiate between treatments.

**Figure 1:**
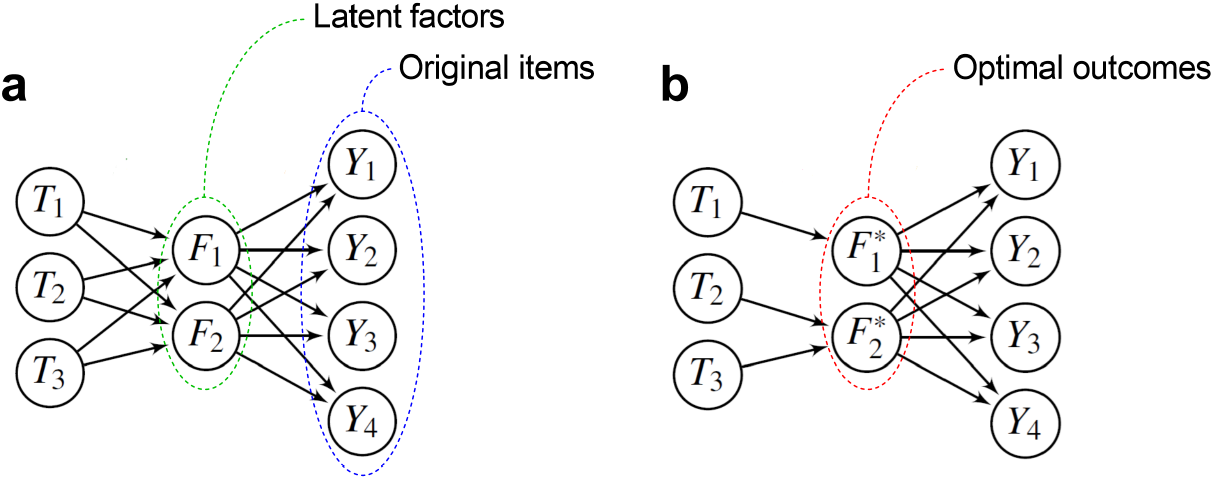
Overview of Supervised Varimax (SV). (a) The algorithm considers a model where treatments affect a set of latent factors ***F***, representing unobserved biopsychosocial processes, in complex ways. The factors in turn affect the items of a clinical rating scale ***Y*** in complex ways. (b) SV organizes the complexity by discovering an alternative set of optimized factors ***F***^∗^, or *optimal outcomes*, such that each treatment affects the optimal outcomes in a simple and unique way.

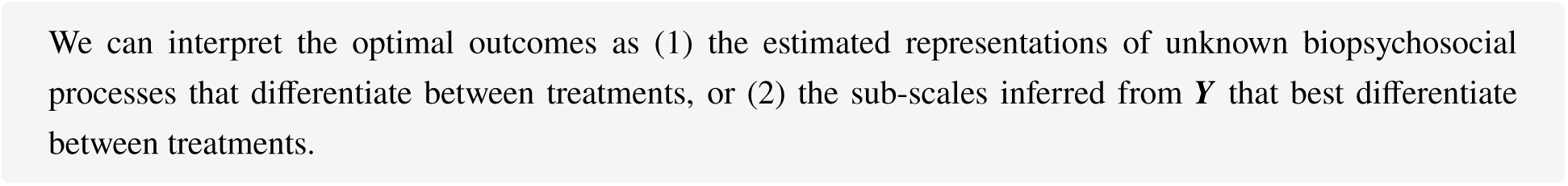

### Independent, Dependent and Nuisance Variables

We set the independent variables as binary treatment assignment, and the dependent variables as the individual items of the original clinical rating scales. We also set age and sex as nuisance variables and therefore partialed out these variables from each rating scale item using ordinary least squares regression before performing downstream analyses.

### Scree Plot with Potentially Meaningful Outcomes

SV learns *m* optimal outcomes by default, where *m* corresponds to the total number of treatments. However, typically only a subset of these outcomes house differential treatment effects large enough to potentially be clinically meaningful. We identify the *potentially meaningful outcomes* by first learning the *m* optimal outcomes using the SV algorithm. We then create a scree plot and eliminate all outcomes at or below the elbow point in the graph. We provide details on how to construct the scree plot in Supplementary Materials 1.6. We only use potentially meaningful outcomes to visualize the output of SV. We also maintain a clear distinction between optimal outcomes that are potentially meaningful and optimal outcomes that are statistically significant, as described below.

### Hypothesis Testing

SV learns the optimal outcomes, so we need to account for the inflated Type I error rate that can result from the estimation process. We thus constructed an omnibus and two post-hoc permutation tests to compute p-values that account for the estimation of optimal outcomes. The null hypothesis of all three permutation tests corresponds to treatment exchangeability and therefore no differential treatment effect. The alternative hypothesis for the omnibus permutation test corresponds to the existence of a differential treatment effect across any of the tested treatments and factors. If we reject the omnibus null hypothesis, then we subsequently perform post-hoc permutation tests of factors, where the alternative hypothesis of each test corresponds to a differential treatment effect in a specific optimal outcome 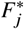. We control for the false discovery rate (FDR) to maximize statistical power. If we finally reject the post-hoc null hypothesis for 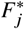, then we perform post-hoc permutation tests of treatment pairs. The alternative hypothesis of each test corresponds to a differential treatment effect between two specific treatments in 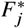. We control for the family-wise error rate (FWER) in this case to guard against even a single false positive. We always perform hypothesis testing using all *m* factors. We provide further details of the omnibus and post-hoc tests in Supplementary Materials 1.7.

### Code Availability

R code for the SV algorithm is available at github.com/ericstrobl/SV.

## Results

### Differentiating Antipsychotics

We first applied SV to Phase I of the CATIE trial, where investigators randomly assigned patients with chronic schizophrenia to five different antipsychotics including: quetiapine, perphenazine, olanzapine, risperidone and ziprasidone. Investigators then tracked the responses of patients using the PANSS score up to 18 months. 1444 patients had complete treatment assignment, age, sex and PANSS item scores at baseline in Phase I. We plot the original Phase I results in Figure 2 (a) after partialing out age and sex as nuisance variables. The CATIE trial suggested that olanzapine is superior to the other antipsychotics by month 18 according to the change in total PANSS score, but this result did not survive multiple comparisons even against ziprasidone [2]. We thus sought to identify optimal outcomes that could differentiate treatment response using the 30 individual items of the PANSS at month 18.

**Figure 2:**
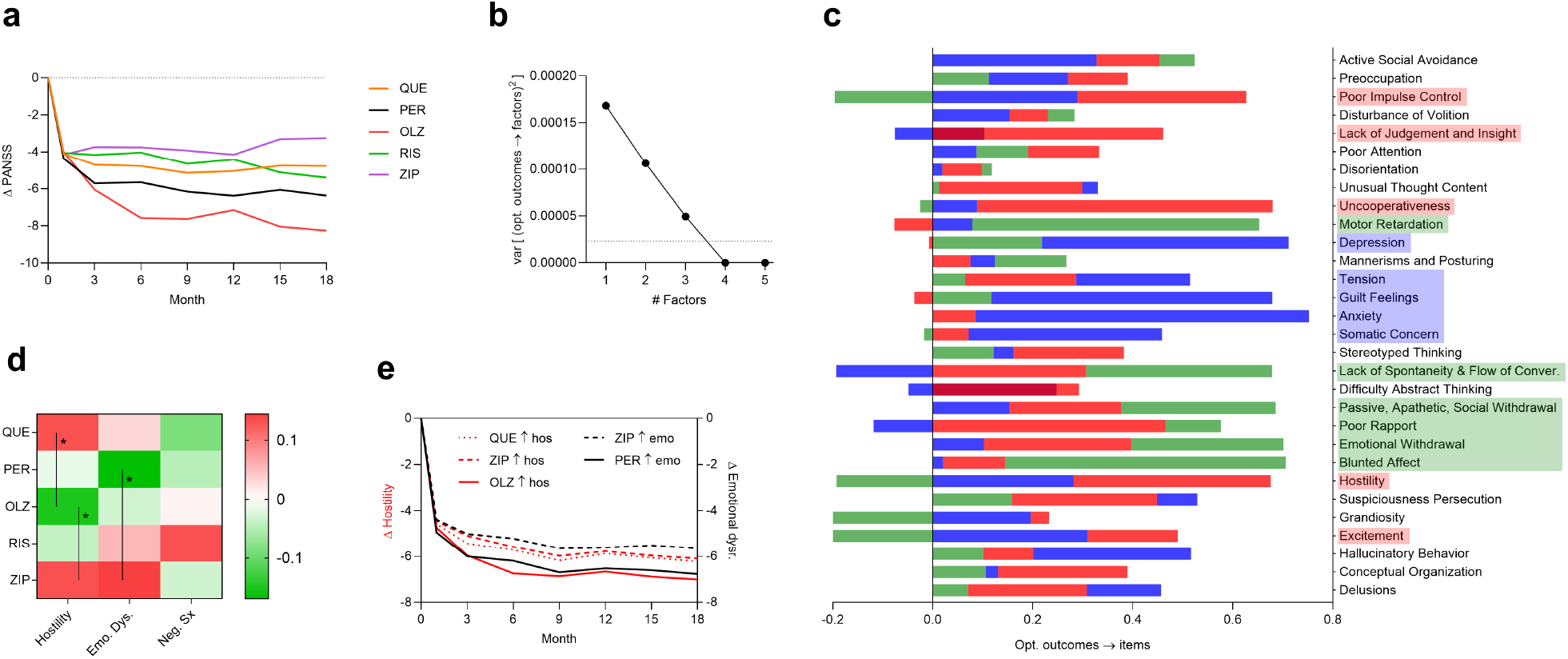
CATIE trial results. (a) The original analysis did not distinguish any particular pair of antipsychotics using the total PANSS score, including olanzapine versus ziprasidone. (b) The scree plot suggested the presence of three potentially meaningful outcomes. (c) Three superimposed bar graphs summarizing the effect sizes from optimal outcomes to individual PANSS items. The potentially meaningful outcomes encapsulated hostility (red), emotional dysregulation (blue) and negative symptoms (green) in chronic schizophrenia. (d) A heatmap of the causal effects from treatments to the three optimal outcomes. Permutation testing revealed that olanzapine is more effective than quetiapine and ziprasidone for hostility. Moreover, perphenazine is more effective than ziprasidone for emotional dysregulation. Asterisks denote treatment pairs with an FWER of less than 0.05. (e) Treating patients with high hostility using olanzapine resulted in a greater reduction in hostility than treating them with quetiapine or ziprasidone (red). Similarly, treating patients with high emotional dysregulation using perphenazine resulted in a greater symptom reduction than ziprasidone (black). QUE denotes quetiapine, PER perphenazine, OLZ olanzapine, RIS risperidone, ZIP ziprasidone.

We ran SV on the CATIE dataset and rejected the omnibus null hypothesis of no differential treatment effects using five optimal outcomes (absolute sum = 1.279, *p* = 0.002). The scree plot of the SV output suggested that only three of the five optimal outcomes were potentially meaningful (Figure 2 (b)). We plot the effect sizes from these three optimal outcomes to individual PANSS items using superimposed bar graphs in Figure 2 (c). The first optimal outcome in red had large positive effects on items related to hostility, such as hostility itself, uncooperativeness, lack of insight and poor impulse control. The second optimal outcome in blue captured emotional dysregulation with large causal effects on items related to low mood and high anxiety. Finally, the third optimal outcome in green involved negative symptoms. The three optimal outcomes thus mapped onto interpretable types of dysfunction seen in schizophrenia. We summarize the effect sizes from treatments to the three optimal outcomes in Figure 2 (d). Note that all treatments had therapeutic effects on all three optimal outcomes. A red cell in Figure 2 (d) does not mean a detrimental or adverse effect on the optimal outcome, but just a worse therapeutic effect than other treatments.

We next tested for specific differential treatment effects. We ran a post-hoc test on each of the five optimal outcomes and rejected the null hypothesis for the first two optimal outcomes corresponding to hostility and emotional dysregulation (absolute sum = 0.469, *p*_*FDR*_ = 0.028; absolute sum = 0.427, *p*_*FDR*_ = 0.037). We finally tested all treatment pairs within the two significant optimal outcomes. Olanzapine had a superior effect on hostility relative to quetiapine and ziprasidone (difference = −0.284, *p*_*FWER*_ = 0.047; difference = −0.283, *p*_*FWER*_ = 0.048). Further, perphenazine had a superior effect on emotional dysregulation relative to ziprasidone (difference = −0.313, *p*_*FWER*_ = 0.020). We conclude that SV identified significant differential treatment effects in two of the five optimal outcomes.

We further tested whether we could translate the above results to everyday clinical practice. Recall that we learned the optimal outcomes using month 18 data, but we wanted to test whether we could predict treatment response like a psychiatrist using insights gained from SV. We therefore derived purposefully simple, non-optimized, non-machine learning based rules with the baseline data. SV discovered that olanzapine is effective for hostility, which we coarsely scored by summing items 7, 22, 26 and 28 of the PANSS. When we give olanzapine to patients with such a hostility score above the median value at baseline, then their hostility score decreases more and faster than patients given quetiapine or ziprasidone (Figure 2 (e) red). Similarly, if we give perphenazine to patients with emotional dysregulation (sum of items 16 and 20) above the median at baseline, then they also improve more and faster than patients given ziprasidone (black). In contrast, the change in total PANSS score in patients with high hostility and high emotional dysregulation does not differ by much from Figure 2 (a) (Supplementary Materials 1.9). We conclude that the insights gained from SV on month 18 data predict treatment response in an appropriate baseline sub-score. Moreover, we can derive simple rules that match clinical common sense: giving patients treatments that best target their given constellation of symptoms improves those symptoms substantially.

### Differentiating Antidepressants

We next sought to identify differential effects of antidepressants in treatment-resistant depression using Level 2 STAR*D data. 1312 patients had complete treatment assignment, age, sex and QIDS-SR item scores in Level 2. The STAR*D dataset is known to be exceptionally challenging, and many investigators have resorted to sophisticated machine learning algorithms in order to accurately match patients better than chance. Visual inspection of the original treatment response curves reveal why – unlike Figure 2 (a), all curves in Figure 3 (a) are near identical.

**Figure 3:**
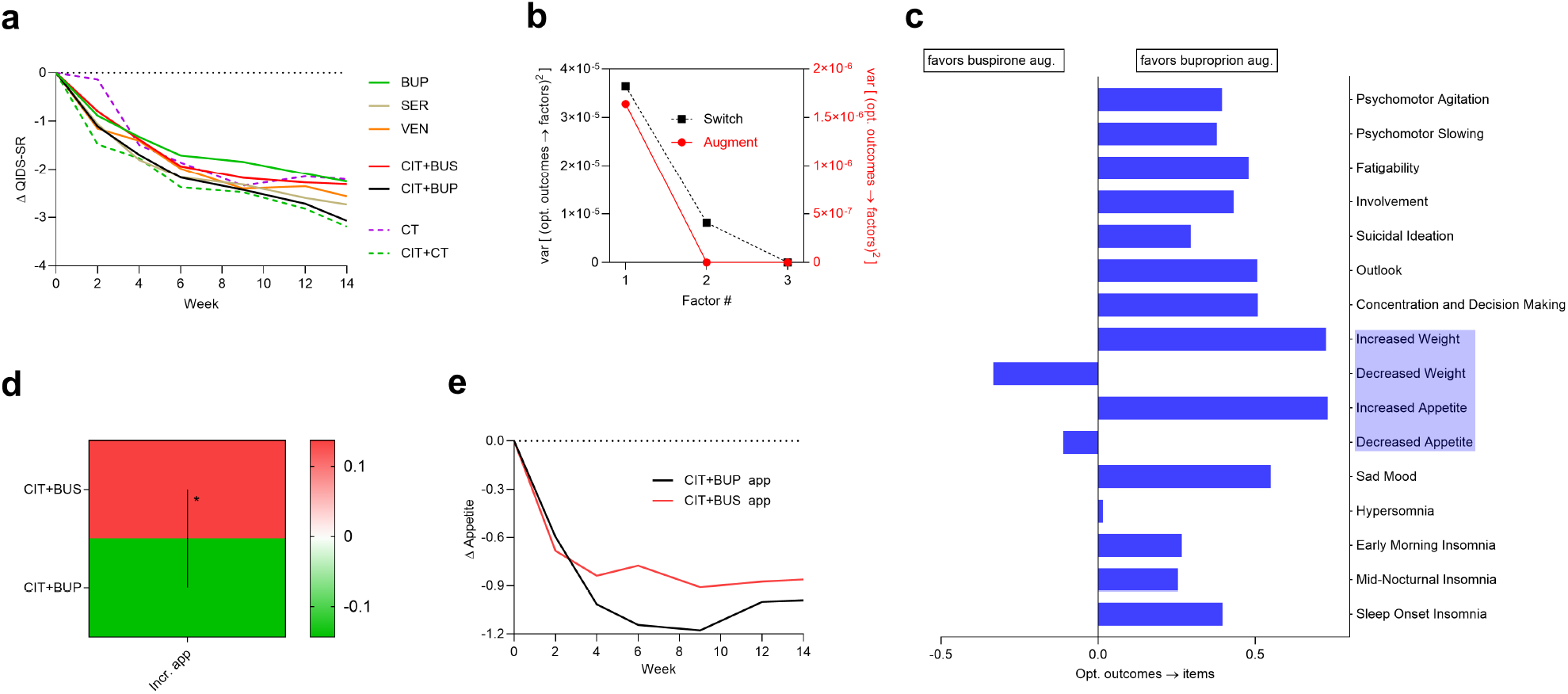
Level 2 STAR*D trial results. (a) All treatments had similar response curves across 14 weeks of treatment. (b) The scree plot suggested the presence of two potentially meaningful outcomes for medication switch and one potentially meaningful outcome for medication augmentation. (c) The one outcome for augmentation corresponded to depression with increased appetite. (d) A heatmap of the effect sizes from treatments to optimal outcomes, where hypothesis testing revealed a significant differential treatment effect between buspirone and bupropion augmentation. (e) A simple clinical rule identified patients with increased appetite at baseline and recapitulated the superior efficacy of bupropion augmentation on appetite relative to buspirone augmentation. Note that bupropion augmentation treats all other non-appetite and non-weight symptoms as well. BUP denotes bupropion sustained release, SER sertraline, VEN venlafaxine extended release, CIT+BUS citalopram with buspirone augmentation, CIT+BUP citalopram with bupropion augmentation, CT cognitive therapy, CIT+CT citalopram with cognitive therapy.

We separately analyzed the switch medications and the augmentation medications where strict randomization took place. We first ran SV on the switch medications, but omnibus testing failed to reject the null (*n* = 659, *p* > 0.05). In contrast, SV detected differential treatment effects among the augmentation medications (*n* = 520, absolute sum = 0.280, *p* = 0.006). The scree plot suggested the presence of one potentially meaningful outcome among the two (Figure 3 (b)). The estimated causal effects from this optimal outcome to the rating scale items corresponded to depression with increased appetite (Figure 3 (c)). Post-hoc testing of treatment pairs resulted in a significant differential effect between buspirone and bupropion augmentation with the one optimal outcome (difference = −0.280, *p*_*FWER*_ = 0.003), Figure 3 (d)). We conclude that bupropion augmentation is particularly effective for patients with treatment-resistant depression and increased appetite.

We next tested the clinical usefulness of the augmentation result by comparing the outcomes of patients with increased appetite (but not bulimia) at baseline who were also randomized to buspirone or bupropion augmentation. We specifically quantified increased appetite by the sum of items 7 and 9 in QIDS-SR. We plot the results in Figure 3 (e). Patients with major depression and increased appetite at baseline who received bupropion augmentation had larger decreases in appetite than patients who received buspirone augmentation. The other symptoms of depression also improved, but the total QIDS-SR score did not consistently capture the differential effect similar to Figure 3 (a) (Supplementary Materials 1.9). We can thus recapitulate the superior effect of bupropion augmentation in major depression with increased appetite using a simple sub-score that identifies patients with increased appetite at baseline.

## Discussion

Total severity scores and remission statuses derived from clinical rating scales are not optimized to differentiate between treatments. We thus introduced the Supervised Varimax (SV) algorithm to optimize the dependent variables instead of the independent ones – just like a psychiatrist differentiates treatments based on their unique *effects* on individual symptoms. SV transforms the individual items of a clinical rating scale into optimal outcomes that maximize differential treatment effect. The algorithm thus can detect subtle differences between the medications, even when the differences are noisily interspersed across multiple individual items. We identified differences in the treatment effects of olanzapine, perphenazine, quetiapine and ziprasidone in chronic schizophrenia. We also identified bupropion augmentation as particularly effective in patients with treatment-resistant depression and increased appetite. Importantly, we detected these differential treatment effects within single RCTs and without any independent variables other than treatment assignment. SV thus does not require deploying, interpreting, generalizing or maintaining a complex machine learning model in the electronic health record.

Note, however, that we do not discount the importance of multiple independent variables. In fact, future work should consider learning the optimal transformation of the independent variables *and* the optimal transformation of the dependent variables in order to maximize predictive performance. We do, nevertheless, claim that *much* more emphasis has been placed on finding transformations of the independent variables rather than on learning the best outcome variables that maximize differential treatment effects. Most existing works only use fixed rating scale scores as the outcome [5, 6], or learn factors/clusters from baseline items in an unsupervised fashion [16, 17, 18]. In this paper, we showed that introducing a supervisory signal from the treatments can substantially improve the learning of outcome measures that differentiate treatments, even without any predictors other than treatment.

The results of SV are congruent with results from large meta-analyses, secondary analyses of adverse effects and clinical intuition. For example, SV identified olanzapine as superior to quetiapine and ziprasidone for hostility in chronic schizophrenia. Two network meta-analyses over tens of thousands of patients have shown that olanzapine has superior efficacy over several antipsychotics in acute agitation in schizophrenia [19, 20]. SV also discovered that perphenazine is particularly effective for emotional dysregulation in chronic schizophrenia; perphenazine is one of the most well-studied first generation antipsychotics in psychotic depression [21]. Furthermore, bupropion augmentation simultaneously treats depression and reduces appetite [22]. SV identified all of these results from only two clinical trials and directly from the rating scales used in the primary analyses. We provide additional technical discussions in Supplementary Materials 1.8.

In summary, existing dependent variables are not optimized to differentiate between treatments. Most investigators have combated this issue by predicting treatment effect using many independent variables in complex machine learning models. However, we can also differentiate between treatments by simply learning dependent variables that achieve such differentiation, such as by the SV algorithm.

## Data Availability

R code is available at github.com/ericstrobl/SV. All datasets are available online at the National Institute of Mental Health (NIMH) Data Archive (https://nda.nih.gov/) with a limited access data use certificate.

## 1. Supplementary Materials (Online Only)

### 1.1. Strategy Overview

We now provide technical details. We use italicized letters like *A* to denote a single random variable, and bold italicized letters like ***A*** to denote a set of random variables. Recall that *orthonormal random variables* have an identity covariance matrix, while *orthonormal parameters* have an identity inner product.

We consider a principal component analysis (PCA) or factor analysis model, where a set of orthonormal factors ***F*** have causal effect sizes *W* on a set of centered items ***Y*** (Figure 4 (a)). Each factor is a linear combination of the items in ***Y*** rather than a cluster. However, unlike traditional PCA or factor analysis, we also consider a third layer, where binary treatments have causal effect sizes *M* on the factors (Figure 4 (b)). The matrices *M* and *W* are both very dense in general.

**Figure 4:**
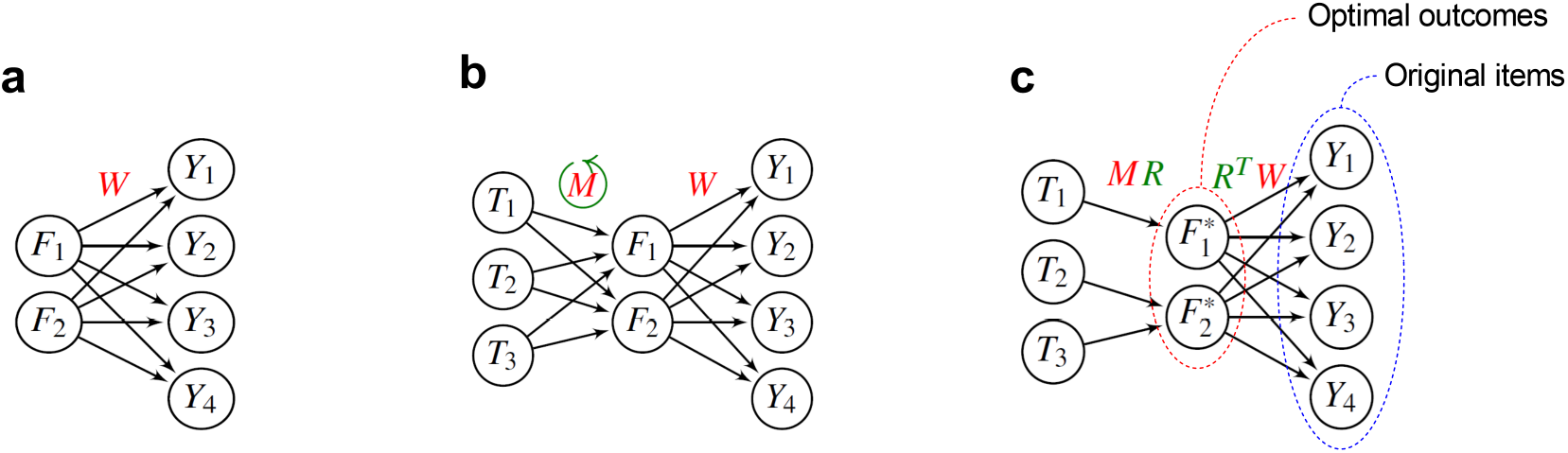
Algorithm overview. (a) The traditional PCA or factor analysis model where factors ***F*** have causal effect sizes *W* on the dependent variables ***Y*** . (b) SV augments the model in (a) using treatments ***T*** with causal effects *M* on ***F***. The algorithm then applies a varimax rotation to *M*. (c) The rotation makes each column of the rotated causal effects *MR* sparse and different. As a result, only a few distinct treatments now cause each factor in ***F***^∗^, or the optimal outcomes.

Supervised Varimax (SV) first learns the set of orthonormal factors ***F*** using PCA. SV then rotates the matrix *M* using the rotation matrix *R* found by Varimax [23] to make each column of *M R* as sparse and as different from the other columns as possible. This ensures that each treatment affects a small and distinct set of rotated factors ***F***\* = ***F****R* (Figure 4 (c)). The rotated factors are still orthonormal and still correspond to linear combinations of the items in ***Y***, but now the factors also maximally differentiate between treatments. We thus also call ***F***\* the *optimal outcomes*.

### 1.2. Model

We describe the underlying model in detail. We consider a supervised component analysis model, where *m* treatment assignments ***T*** causally affect *q* orthonormal factors ***F*** that in turn cause *p* dependent variables ***Y*** (Figure 4 (b)). We assume that ***Y*** is centered to expectation zero. The treatments causally affect the dependent variables ***Y*** as represented by the following structural equation:

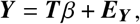

where ***E***_***Y***_ denotes a vector of independent and identically distributed (i.i.d.) error terms with mean zero and covariance Σ. The error terms do not necessarily follow a Gaussian distribution, and Σ may have non-zero off-diagonal elements. Let *V*Λ*V*^*T*^ correspond to the eigendecomposition of the covariance matrix of ***Y***, equivalent to PCA of ***Y***, where *V* is a *p* × *p* matrix of *p* eigenvectors and Λ denotes a diagonal matrix of *p* strictly positive eigenvalues. The set ***Y*** may contain many correlated variables, which we can transform into a set of *q* ≤ *p* orthonormal ones using the eigendecomposition:

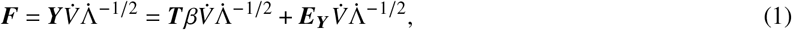

where 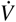 refers to *q* eigenvectors associated with (not necessarily the largest) *q* eigenvalues in the diagonal entries of 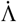 . Now let 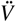 and 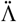 denote the remaining *p* − *q* eigenvectors and their associated eigenvalues, respectively. We set 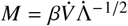 and 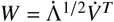, so that we recover the following model depicted in Figure 4 (b):

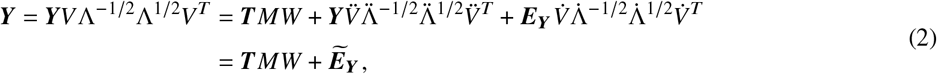

where 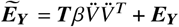.

### 1.3. Optimal Rotation

The compression to *q* factors invariably results in some loss of information of the causal effect from ***T*** to ***Y*** as seen in 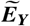 of Equation (2). However, the transformation matrix 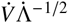 in Equation (1) is not unique because the columns of ***F****R* are orthonormal as well, where *R* corresponds to a rotation matrix. We can thus also take advantage of the orthonormality by choosing a rotation that maximally differentiates treatment effects on the set of factors:

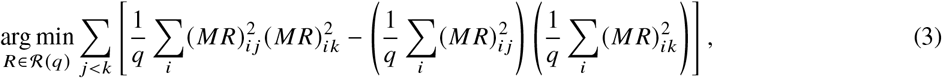

where ℛ (*q*) denotes the set of all *q* × *q* rotation matrices. In other words, we minimize the covariance of pairs of columns of treatment effects squared, and then sum over all possible pairs. As a result, each factor tends to be caused by a different set of treatments.

The minimization problem in Expression (3) yields the same solution as the following maximization problem, also known as the varimax rotation [23]:

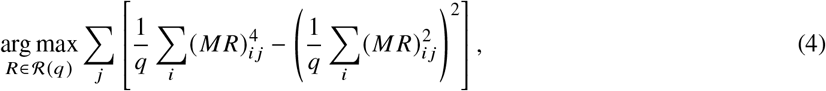

where we maximize the variance of each column of (*M R*) ^(2)^ (element-wise squared). In other words, we maximize a quantity similar but not equivalent to the kurtosis of each column of *M R* to induce outliers and sparsity. We combine Expressions (3) and (4) to see the equivalence between the maximization and minimization as follows:

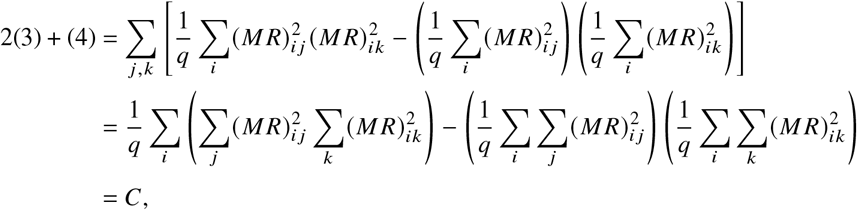

which is equal to a constant *C* because rotations preserve row vector lengths, or the sum of squares of its elements. Hence, the minimization problem in Expression (3) yields the same solution as the maximization problem in Expression (4). If *R* is the solution to Expression (4), then we refer to ***F***^∗^ = ***F****R* as the *optimal outcomes*.

Varimax is known to approximately induce part of Thurstone’s *simple structure* [24] in *M R*, which we paraphrase for the present context below:

1. each factor has no causal effect from most treatments;
2. each factor has large causal effects, in magnitude, from a small number of treatments;
3. few factors have large causal effects, in magnitude, from the same treatment.

In particular, the row sums of the squared elements of *M R* remain fixed under rotation because *R* is orthonormal. We thus maximize the variance depicted in Expression (4), or the mean of squared pairwise distances, so that most squared elements of *M R* are large or zero; this in turn satisfies the first two items listed above. The third follows by equivalently minimizing the covariance shown in Expression (3).

### 1.4. Supervised Varimax

We are now ready to describe the proposed algorithm, which we summarize in Algorithm 1. SV first standardizes ***Y*** and then performs an eigendecomposition of the correlation matrix of ***Y***. The algorithm extracts the eigenvectors associated with the top *q* largest eigenvalues in Line 2. We set *q* = *m* as a default value, where *m* denotes the cardinality of ***T***. We now have 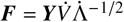 corresponding to the unrotated factors. SV then regresses ***F*** on ***T*** to obtain the causal effects *M* in Line 3. Next, SV sparsifies *M* with a varimax rotation in order to compute the optimal outcomes ***F***^∗^ in Lines 4 and 5, respectively. Note that Varimax has permutation and sign indeterminancies [25], which we determine in Line 6 by sorting ***F***^∗^ according to variance explained and non-negatively correlating ***F***^∗^ to Σ _*k*_ *Y*_*k*_ via sign flips. SV thus ultimately outputs the desired matrices *M R, R*^*T*^*W* and ***F***^∗^ with permutation and sign determinancy.

#### Algorithm 1

**Supervised Varimax**

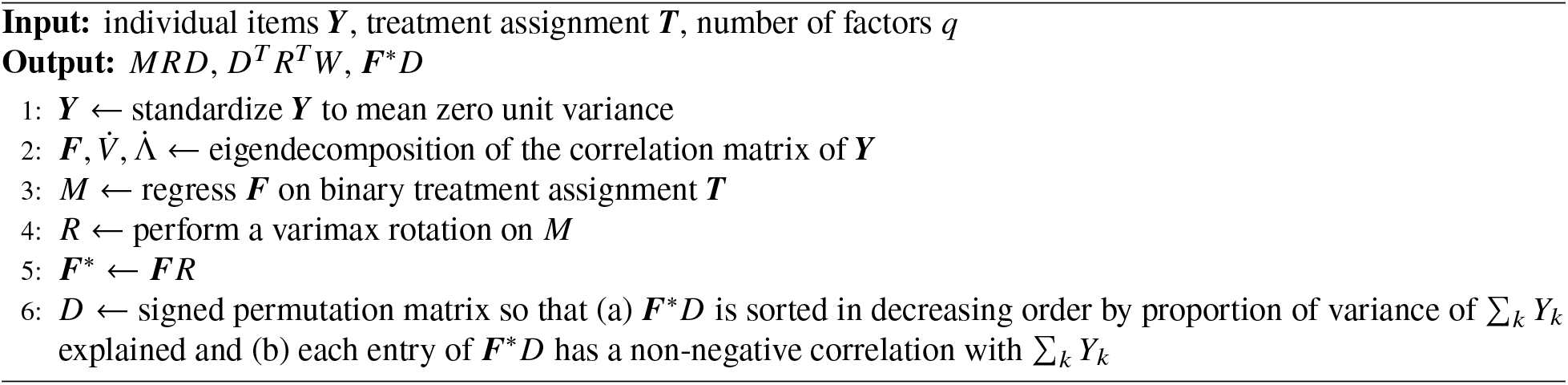

### 1.5. Empirical Performance

#### 1.5.1. Synthetic Data

We assessed the performance of SV by comparing it against alternative algorithms, including PCA [26], PCA with Varimax (PCA+VM) [25], factor analysis with Varimax (FA+VM) [23] and independent component analysis (ICA) [27]. We first generated synthetic data by drawing 1000 samples from the model shown in Equation (2), where each entry of ***E***_***Y***_ followed an independent t-distribution with three degrees of freedom; we chose this non-Gaussian distribution to ensure identifiability of the ICA solution. We sampled the matrices *M* and *W* by drawing each entry from Unif([−1, −0.25] ∪ [0.25, 1]). We then performed a varimax rotation on the ground truth matrix *M* to yield the rotation matrix *R*. We removed sign and permutation indeterminancies using the same procedure as Line 6 in SV. We repeated the above process 1000 times for 2, 3 and 4 factors in ***F***. We thus generated a total of 3 × 1000 = 3000 unique datasets.

We summarize the results in Figure 5. Recall that SV maximizes the sum of the variances of the columns of (*M R*) ^(2)^ via Expression (4), where a higher value corresponds to increased sparsity. SV correspondingly identified the sparsest matrix *M R* as assessed by the mean of those variances (Figure 5 (a)). SV estimated sparser matrices with more factors, whereas other methods did not display this improvement. The algorithm also estimated the matrices *M R* and *R*^*T*^*W* with the lowest root mean square error (RMSE) to their ground truths (Figures 5 (b) and (c)). Each of these three comparisons held at a Bonferonni corrected threshold of 0.05/4 according to paired t-tests, since we compared SV against a total of four other algorithms. We conclude that SV outperformed all other algorithms.

**Figure 5:**
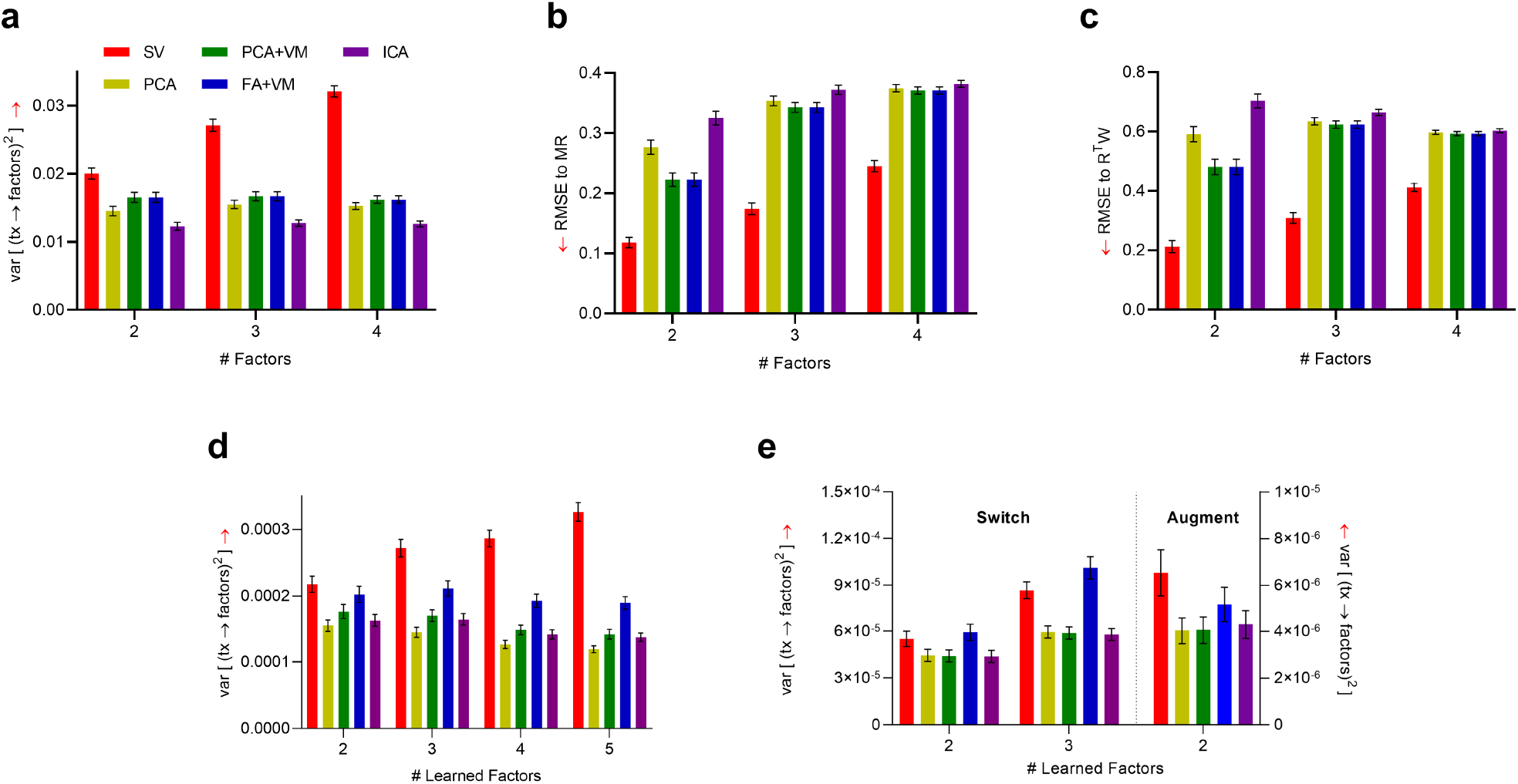
Empirical performance results. (a) SV identified the sparsest matrix *MR* across 1000 simulated models. SV also computed the matrix *MR* and the causal effects *R*^*T*^ *W* from factors to ***Y*** with the highest accuracy in (b) and (c), respectively. (d) SV estimated the sparsest matrix *MR* in the CATIE Phase I dataset. (e) The algorithm also estimated the sparsest matrix among the augmentation medications in Level 2 of STAR*D, but not among the switch medications. Error bars denote 95% confidence intervals of the mean. Red arrows denote direction of better performance.

#### 1.5.2. Real Data

We tested whether SV outperformed existing algorithms in Phase I of CATIE by running each algorithm on 1000 bootstrap datasets. We do not have access to the ground truth matrices of *M R* and *R*^*T*^*W* in real data. We thus assessed how well the algorithms differentiate between treatments by computing the mean variance across the columns of (*M R*) ^(2)^ similar to Figure 5 (a). We found that SV achieved the largest variance with all learned numbers of factors (Figure 5 (d)). Further, the performance of SV only continued to improve with an increasing number of learned factors, whereas other algorithms plateaued. We conclude that SV estimated the sparsest matrix *M R* in the CATIE dataset, and the results of SV mimicked those seen with the synthetic data.

We next ran SV on the switch medications in Level 2 of STAR*D. Unfortunately, factor analysis with Varimax estimated a sparser matrix *M R* than SV according to the mean variance of the columns of (*M R*) ^(2)^ across all numbers of learned factors (Figure 5 (e)). SV, however, outperformed all other algorithms in estimating a sparser *M R* among the augmentation medications, including factor analysis with Varimax with only two learned factors (paired t-test, *t* = 12.11, *p* < 0.001). We conclude that SV outperformed all other algorithms for the augmentation medications only. Recall that these results are consistent with the omnibus and post-hoc permutation tests, where SV detected significant differential treatment effects between the augmentation but not switch medications.

#### 1.6. Potentially Meaningful Outcomes and Scree Plot

We identify the *potentially meaningful outcomes* by first learning the *q* = *m* optimal outcomes using the SV algorithm. We then plot the optimal outcomes against the variances of their columns in (*M R*) ^(2)^ . We plot the variances in decreasing order. We then eliminate all factors at and below the elbow point of the resultant graph, which we find to be very close to zero in practice. For example, we eliminate the optimal outcomes associated with the fourth and fifth smallest variances in Figure 2 (b). We usually retain only a small number of optimal outcomes after this diagnostic step.

### 1.7. Permutation Tests

#### 1.7.1. Omnibus Test

We consider the following omnibus null and alternative hypotheses:

- ℋ_0_ : treatments are exchangeable so that no differential treatment effect exists;
- ℋ_1_ : a differential treatment effect exists for some factor.

We operationalize the above omnibus hypotheses as follows:

- ℋ_0_ : ***Y* ╨** *T*,
- ℋ_1_ : ∑_*ij*_| (*M R*)_*ij*_ | >(∑_*ij*_ | (*M R*)_*ij*_ |)_***Y* ╨***T*_ .

We call ∑_*i j*_ | (*M R*)_*ij*_ | the *absolute sum*, and the notation (∑ _*i j*_ | (*M R*)_*ij*_ |(_***Y* ╨***T*_ refers to the absolute sum when the null hypothesis holds. We permute treatment assignment, run SV, and then compute the absolute sum in each permutation. We finally count the proportion of cases where the statistic falls at or above the same quantity computed on the original samples after 100,000 permutations.

#### 1.7.2. Post-Hoc Test for Factors

If we reject the omnibus null hypothesis, then we test each factor using the following post-hoc hypotheses:

- ℋ_0_ : treatments are exchangeable so that no differential treatment effect exists;
- ℋ_1_ : a differential treatment effect exists for some factor 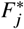.

We operationalize these hypotheses as:

- ℋ_0_ : ***Y* ╨** *T*,
- ℋ_1_ : Σ_*i*_ | (*M R*)_*i j*_ | > (Σ_*i*_ | (*M R*)_*ij*_ | _***Y* ╨***T*_,

where we now have only summed over the treatments in the absolute sum statistic. We again permute treatment assignment, run SV, and then compute the absolute sum for 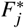 on each permuted sample. We finally count the proportion of cases where the statistic falls at or above the same quantity computed on the original samples after 100,000 permutations. Repeating the above procedure for each factor leads to a vector of *q* p-values. Note that we posit the existence of multiple optimal outcomes and thus seek to detect optimal outcomes with high statistical power at the expense of a few false positives, rather than guard against even a single false positive. We thus test all *q* factors. We simultaneously estimate the false discovery rate (FDR) using the permutation-based technique described in [28].

#### 1.7.3. Post-Hoc Test for Treatment Pairs

If we reject the above post-hoc null hypothesis for a particular factor 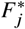 after correcting for multiple comparisons, then we test each pair of treatments *T*_*i*_ and *T*_*k*_ within 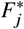 using the following additional post-hoc hypotheses:

- ℋ_0_ : all treatments are exchangeable so that no differential treatment effect exists;
- ℋ_1_ : a differential treatment effect exists between treatments *T*_*i*_ and *T*_*k*_ in factor 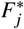.

We wish to guard against even a single false positive in this test because the optimal outcomes may contain a few false positives with only FDR control. We thus mimic Tukey’s range test [29] with the maxT method [30, 31] in order to control the family-wise error rate (FWER) among all possible treatment pairs within an optimal outcome:

- ℋ_0_ : ***Y* ╨** *T*,
- ℋ_1_ : | (*M R*)_*ij*_ − (*M R*)_*kj*_ | > max_*i*_ (*M R*)_*ij*_ − min_*i*_ (*M R*)_*ij* ***Y* ╨***T*_,

where max_*i*_ (*M R*)_*ij*_ − min_*i*_ (*M R*)_*ij*_ corresponds to the *range*. We also call | (*M R*)_*ij*_ − (*M R*)_*kj*_ | the *absolute difference* statistic, and (*M R*)_*ij*_ − (*M R*)_*kj*_ the *difference* statistic. We permute treatment assignment, run SV, and then compute the range statistic in each permutation. We finally count the proportion of cases where the range falls at or above the absolute difference computed on the original samples after 100,000 permutations.

### 1.8. Technical Discussion

SV achieves high statistical power because it effectively models the heterogeneity of mental illnesses. The algorithm deconvolutes mental illness into a small number of latent factors that optimally combine all items, rather than cluster items into disjoint categories. The factors likely correspond to complex biopsychosocial processes that can only be approximately gleaned from existing clinical rating scales. Moreover, SV represents each patient as a weighted combination of these complex processes, rather than as a single category or biotype.

SV does not seek to maximize probabilistic independence between factors like ICA [27]. Instead, the algorithm only identifies orthogonal factors, or independence up to the second moment. We are not interested in identifying factors that satisfy all mathematical constraints associated with independence, but only enough constraints so that the factors correspond to roughly distinct biopsychosocial processes that lead to clinically actionable insights. SV thus leverages the additional rotational indeterminancy of orthogonality to identify differential treatment effects rather than maximize independence.

We must temper the above strengths with some limitations. SV imposes a linear model, even though the effect from medications to items may depend non-linearly on the dose of each medication. The algorithm also sets the number of factors *q* to the number of treatments *m* as a heuristic, but we may want to select *q* in a data-driven manner with subsequent post-model selection inference. Third, we limited the present study to pre-specified diagnoses and associated rating scales, even though increased diversity in the dependent variables can introduce more degrees of freedom to differentiate treatment effects. Finally, SV currently requires data from randomized clinical trials, but the algorithm may benefit substantially from the diversity and large sample sizes seen in observational data with proper confounder control. Future work should therefore investigate multiple scales across multiple mental illnesses by modifying SV to perform well with observational data.

### 1.9. Change in Total Scores

**Figure 6:**
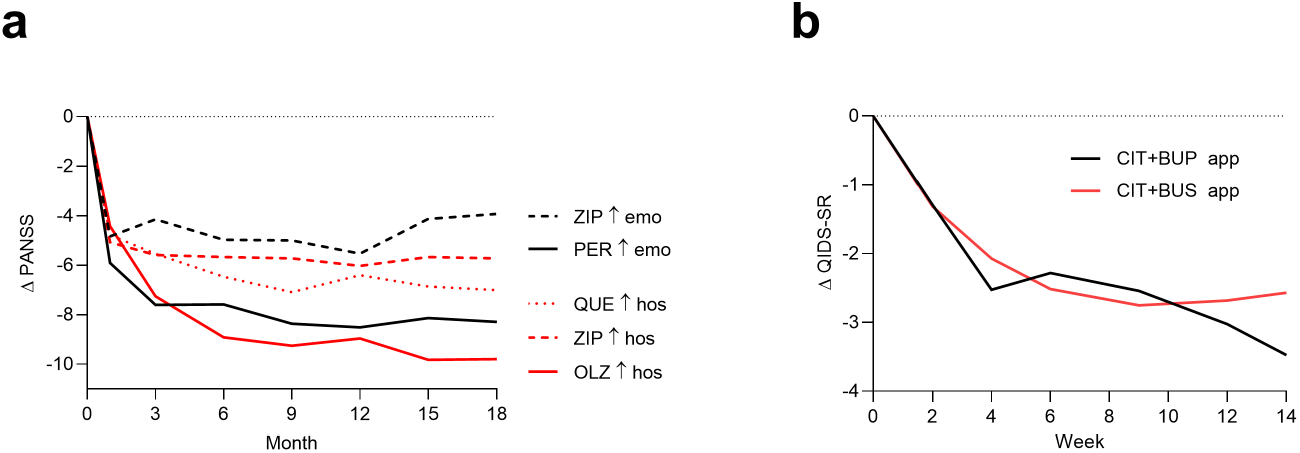
Total severity scores do not effectively differentiate between treatments. As a result, changes in the total score of PANSS in (a) and QIDS-SR in (b) simply mimic those seen in Figures 2 (a) and 3 (a), respectively – even in the identified subpopulations.

## References

[1] Fernandes BS, Williams LM, Steiner J, Leboyer M, Carvalho AF, Berk M. The new field of ‘precision psychiatry’. BMC Medicine. 2017;15:1–7.

[2] Lieberman JA, Stroup TS, McEvoy JP, Swartz MS, Rosenheck RA, Perkins DO, et al. Effectiveness of antipsychotic drugs in patients with chronic schizophrenia. New England Journal of Medicine. 2005;353(12):1209–23.

[3] Trivedi MH, Fava M, Wisniewski SR, Thase ME, Quitkin F, Warden D, et al. Medication augmentation after the failure of SSRIs for depression. New England Journal of Medicine. 2006;354(12):1243–52.

[4] Rush AJ, Trivedi MH, Wisniewski SR, Stewart JW, Nierenberg AA, Thase ME, et al. Bupropion-SR, sertraline, or venlafaxine-XR after failure of SSRIs for depression. New England Journal of Medicine. 2006;354(12):1231–42.

[5] Chekroud AM, Zotti RJ, Shehzad Z, Gueorguieva R, Johnson MK, Trivedi MH, et al. Cross-trial prediction of treatment outcome in depression: a machine learning approach. The Lancet Psychiatry. 2016;3(3):243–50.

[6] Nie Z, Vairavan S, Narayan VA, Ye J, Li QS. Predictive modeling of treatment resistant depression using data from STAR* D and an independent clinical study. PloS One. 2018;13(6):e0197268.

[7] Iniesta R, Malki K, Maier W, Rietschel M, Mors O, Hauser J, et al. Combining clinical variables to optimize prediction of antidepressant treatment outcomes. Journal of Psychiatric Research. 2016;78:94–102.

[8] Iniesta R, Hodgson K, Stahl D, Malki K, Maier W, Rietschel M, et al. Antidepressant drug-specific prediction of depression treatment outcomes from genetic and clinical variables. Scientific Reports. 2018;8(1):1–9.

[9] Koutsouleris N, Kahn RS, Chekroud AM, Leucht S, Falkai P, Wobrock T, et al. Multisite prediction of 4-week and 52-week treatment outcomes in patients with first-episode psychosis: a machine learning approach. The Lancet Psychiatry. 2016;3(10):935–46.

[10] Leighton SP, Upthegrove R, Krishnadas R, Benros ME, Broome MR, Gkoutos GV, et al. Development and validation of multivariable prediction models of remission, recovery, and quality of life outcomes in people with first episode psychosis: a machine learning approach. The Lancet Digital Health. 2019;1(6):e261–70.

[11] Jaworska N, De la Salle S, Ibrahim MH, Blier P, Knott V. Leveraging machine learning approaches for predicting antidepressant treatment response using electroencephalography (EEG) and clinical data. Frontiers in psychiatry. 2019;9:768.

[12] Chekroud AM, Hawrilenko M, Loho H, Bondar J, Gueorguieva R, Hasan A, et al. Illusory generalizability of clinical prediction models. Science. 2024;383(6679):164–7.

[13] Falkai P, Koutsouleris N. Why is it so difficult to implement precision psychiatry into clinical care? The Lancet Regional Health–Europe. 2024;43.

[14] Kay SR, Fiszbein A, Opler LA. The positive and negative syndrome scale (PANSS) for schizophrenia. Schizophrenia Bulletin. 1987;13(2):261–76.

[15] Rush AJ, Trivedi MH, Ibrahim HM, Carmody TJ, Arnow B, Klein DN, et al. The 16-Item Quick Inventory of Depressive Symptomatology (QIDS), clinician rating (QIDS-C), and self-report (QIDS-SR): a psychometric evaluation in patients with chronic major depression. Biological Psychiatry. 2003;54(5):573–83.

[16] Chekroud AM, Gueorguieva R, Krumholz HM, Trivedi MH, Krystal JH, McCarthy G. Reevaluating the efficacy and predictability of antidepressant treatments: a symptom clustering approach. JAMA psychiatry. 2017;74(4):370–8.

[17] Collins KA, Eng GK, Tural Ü, Irvin MK, Iosifescu DV, Stern ER. Affective and somatic symptom clusters in depression and their relationship to treatment outcomes in the STAR*D sample. Journal of Affective Disorders. 2022;300:469–73.

[18] Silverstein B, Patel P. Poor response to antidepressant medication of patients with depression accompanied by somatic symptomatology in the STAR* D Study. Psychiatry Research. 2011;187(1-2):121–4.

[19] Huhn M, Nikolakopoulou A, Schneider-Thoma J, Krause M, Samara M, Peter N, et al. Comparative efficacy and tolerability of 32 oral antipsychotics for the acute treatment of adults with multi-episode schizophrenia: a systematic review and network meta-analysis. The Lancet. 2019;394(10202):939–51.

[20] Paris G, Bighelli I, Deste G, Siafis S, Schneider-Thoma J, Zhu Y, et al. Short-acting intramuscular second-generation antipsychotic drugs for acutely agitated patients with schizophrenia spectrum disorders. A systematic review and network meta-analysis. Schizophrenia Research. 2021;229:3–11.

[21] Oliva V, Possidente C, De Prisco M, Fico G, Anmella G, Hidalgo-Mazzei D, et al. Pharmacological treatments for psychotic depression: a systematic review and network meta-analysis. The Lancet Psychiatry. 2024;11(3):210–20.

[22] Mohamed S, Johnson GR, Chen P, Hicks PB, Davis LL, Yoon J, et al. Effect of antidepressant switching vs augmentation on remission among patients with major depressive disorder unresponsive to antidepressant treatment: the VAST-D randomized clinical trial. JAMA. 2017;318(2):132–45.

[23] Kaiser HF. The varimax criterion for analytic rotation in factor analysis. Psychometrika. 1958;23(3):187–200.

[24] Thurstone LL. Multiple-factor Analysis: A Development and Expansion of The Vectors of the Mind. University of Chicago Committee on Publications in Biology and Medicine. Publications. University of Chicago Press; 1947.

[25] Rohe K, Zeng M. Vintage factor analysis with Varimax performs statistical inference. Journal of the Royal Statistical Society Series B: Statistical Methodology. 2023 07;85(4):1037–60.

[26] Pearson K. On lines and planes of closest fit to systems of points in space. The London, Edinburgh, and Dublin Philosophical Magazine and Journal of Science. 1901;2(11):559–72.

[27] Lee TW, Lee TW. Independent component analysis. Springer; 1998.

[28] Xie Y, Pan W, Khodursky AB. A note on using permutation-based false discovery rate estimates to compare different analysis methods for microarray data. Bioinformatics. 2005;21(23):4280–8.

[29] Tukey JW. Comparing individual means in the analysis of variance. Biometrics. 1949:99–114.

[30] Westfall P. Resampling-Based Multiple Testing: Examples and Methods for p-Value Adjustment. Wiley; 1993.

[31] Rempala GA, Yang Y. On permutation procedures for strong control in multiple testing with gene expression data. Statistics and its Interface. 2013;6(1).

